# Physical Activity Levels and Associated Factors among Upper Primary School Children in Lusaka, Zambia: Implications for Health Interventions

**DOI:** 10.64898/2026.04.17.26351077

**Authors:** Simon Himalowa, John Zulu, Titus Haakonde, Joseph Lupenga, Richard Kunda, Yvonne Colgrove, José Frantz, Margaret M. Mweshi, Martha Banda

**Author notes:** **Corresponding Author:** Simon Himalowa.

## Abstract

**Introduction:** Physical inactivity and sedentary behaviour are significant risk factors for non-communicable diseases. Engaging in regular physical activity (PA) during childhood is crucial for preventing long-term health burdens. This study examined PA levels and associated factors among upper primary school children in Lusaka, Zambia.

**Methodology:** A cross-sectional survey was conducted from August to October 2022 among 638 children aged 9–18 years from six public and six private schools. Data were collected using the Physical Activity Questionnaire for Children (PAQ-C), Youth Risk Behaviour Survey (YRBS), Model of Youth Physical Activity Questionnaire (MYPA), and 3-Day Physical Activity Recall Questionnaire (3DPAR). Analyses included descriptive statistics, Chi-square, Fisher’s exact tests and multivariable binary logistic regression at a 0.05 significance level and 95% confidence interval.

**Results:** Most participants (82%) were insufficiently active, with only 18% achieving sufficient PA. Reported barriers included lack of playgrounds or parks near home (p=0.012), neighbourhood safety concerns (p=0.041), and limited parental supervision (p=0.006). Watching television reduced the odds of PA by 69% (aOR=0.31; 95% CI: 0.13-0.75). Conversely, peer support increased activity by 15% (aOR=1.15, 95% CI: 0.67–1.97), while not being concerned about showering or fixing hair after PA increased activity by 94% (aOR=1.94; 95% CI: 1.21–3.11).

**Conclusion:** The majority of school children in this study did not meet recommended PA levels. Barriers to activity included personal, parental, and environmental factors. Interventions should prioritise safe play spaces, increased parental and peer support, and reduced screen time to curb future non-communicable disease risks.

## Background

Physical activity, defined as any skeletal muscle movement that requires energy expenditure, is a cornerstone of child and adolescent health [1]. The health benefits of PA include improved cardiorespiratory, muscular fitness and bone health, improved mental health, quality of life and well-being and reduced risks of non-communicable diseases (NCDs) [1,2]. The World Health Organization (WHO) recommends that children aged 5-17 years engage in at least 60 minutes of moderate- to vigorous-intensity PA daily that is mostly aerobic [1,2]. The activities should incorporate those that strengthen muscle and bone, at least three (3) times per week [1,2]. However, global trends indicate that over 70% of adolescents (ages 11–17) fail to meet the WHO minimum recommended PA levels [2]. This growing inactivity is often attributed to increased screen time, reduced opportunities for active transport, and limited safe recreational spaces, resulting in sedentary lifestyles that track into adulthood [3].

In recent years, research across Africa has shown similar trends, with only 20% in-school adolescents meeting the WHO’s recommended physical activity levels, though with variations across countries [4]. Studies indicate that South African adolescents experience rising sedentary behaviours [5], while children in Benin report comparatively higher PA levels, partly due to lower urbanisation and reliance on active commuting [4]. School settings play a central role, as they provide structured opportunities for PA through physical education, recess, and after-school activities [6,7]. However, even where policies exist, implementation is often inconsistent, and cultural, infrastructural, and socioeconomic barriers persist [7–9].

In Zambia, national data on PA among school-aged children remain limited. The country’s urbanisation trends and technological advancements have led to the confinement of children indoors [10] and increased sedentary lifestyles. The lack of structured PA programs, excessive screen time, and inadequate recreational spaces have further contributed to physical inactivity. Physical inactivity in childhood has been linked to adverse health outcomes, including increased obesity and reduced exercise tolerance, as well as diminished academic performance due to decreased mental acuity [6,11,12]. Therefore, this study assessed the physical activity levels and associated factors among upper primary school children in Lusaka.

## Methods

### Study Design and Setting

A cross-sectional survey was conducted in 12 schools (6 public and 6 private) in Lusaka Urban District from August to October, 2022. The selection of these schools ensured representation across different socioeconomic backgrounds.

### Population and sampling

A total of 2, 579 school-going children attending upper primary (grades five to seven) from all the 10 zones in Lusaka Urban District were considered 6 zones out of the 10 were randomly selected to give all the 10 zones an equal chance of being selected. Six schools were included, one from each of the 6 selected zones, to give this study a better statistical inference. All the names of the 10 zones were put in plain envelopes and placed in a bowl, where 6 zones were randomly selected using the fishbowl method [13]. From the 6 selected zones, 6 schools were then randomly selected and included in the study. Because private schools are not evenly mapped, purposeful sampling was used to select 6 private schools evenly matched for socioeconomic status. One thousand nine hundred and sixty-six (1, 966) children were from public schools and 613 were from private schools. According to the Yamane formula [14], the minimum sample size needed for public schools was 332, while that of private schools was 242. The Yamane formula is popular for cross-sectional research because of its simplicity, efficiency, and accuracy in determining an adequate sample size, ensuring valid results without high costs or effort. The dropout rate was factored in and was calculated at 10%, bringing the total sample size of public schools to 369 and that of private schools to 269, yielding a total of 638 participants.

### Data Collection and Instrument

Data were collected from August to October, 2022 using a self-administered standardised questionnaire, with assistance provided by the researcher and trained assistants when needed. Informed consent was obtained from participants, with assent from parents or guardians and school authorities. The questionnaire, scored on a 5-point Likert scale, was adapted from four instruments: the Physical Activity Questionnaire for Children (PAQ-C) to assess levels of physical activity and influencing factors [15]; selected items (questions 43 and 44) from the Youth Risk Behaviour Survey (YRBS) to capture sedentary behaviour such as screen time (YRBS, 2019); the Model of Youth Physical Activity Questionnaire (MYPA) to examine factors including parental support, peer support, self-efficacy, enjoyment, and social barriers [16]; and the 3-Day Physical Activity Recall Questionnaire (3DPAR) to evaluate environmental barriers related to equipment access and neighbourhood safety [17].

For this study, children who engaged in PA two (2) times or less a week were considered insufficiently active, those who engaged in PA three (3) to four (4) times were considered to be moderately active, those who were active five (5) to six (6) times were considered to be sufficiently active while those that engaged in PA seven (7) or more times were considered to be highly active.

This is a valid and reliable question assessing school children or adolescents’ PA at an epidemiological level [3], and it has been used in several studies [3,18]. School children were provided with a definition of PA accompanied by examples of some age-relevant activities (e.g., running, cycling, dancing, playing football, basketball). The PA levels among children were self-reported, as self-report measures remain the most practical and commonly used method of assessing physical activity in epidemiological studies and national surveillance systems [19].

### Study Variables

The outcome variable was the level of physical activity measured on a dichotomous scale (sufficiently active or insufficiently active) [20]. Children who engaged in PA five (5) times or more a week were considered to be sufficiently active, while those who engaged in PA less than five (5) times a week were considered insufficiently active [3,15]. The independent variables included: age, sex, BMI, waist circumference, Parents’/guardians’ level of education, types of school, hours spent watching TV, hours spent playing computer or video games, personal barriers, physical and environmental barriers, parental support and peer support.

### Piloting

The questionnaire was piloted on 10 children at Olive Park, a mid-fee tier private school in Lusaka, which was excluded from the main study. The pilot gave an idea of how long it took to complete the questionnaire and helped to refine clarity and flow. Ultimately, redundant questions were removed from the completed questionnaire. Using standardised instruments ensured consistent data collection on factors influencing children’s participation in PA.

### Statistical analysis

The quantitative data were analysed using STATA version 14 (StataCorp, 2014). All questionnaires were checked for completeness before analysis. Descriptively, continuous variables were summarised using the mean and standard deviation if the data met the normality assumption; otherwise, the median and interquartile range were used. Categorical variables were presented using frequencies and percentages. Cross tabulations were tested using the Chi-square and Fisher’s exact test. A multivariable logistic regression was used to identify risk factors associated with the level of physical activity, reporting the odds ratio and their 95% confidence interval. All the results were set to be significant at the P<0.05 level.

## Results

### Demographic Characteristics

The study included 638 upper primary school children. The mean age of the participants was 12 years (SD± 1.6). The majority of the participants were females (54%), had normal BMI (53%), and were from public schools (58%). The highest level of education attained by the mothers, fathers and guardians of the children was tertiary education (43%, 49% and 40% respectively). The prevalence of sufficient PA was 18.18% (95% CI: 15.26-21.40). The rate of sufficient PA was significantly high among children whose mothers had no formal education (52.0%), those who did not watch TV (26.47%), and those who spent more than five hours playing computer or video games (39.62%) (p<0.05) (Table 1).

**Table 1:**
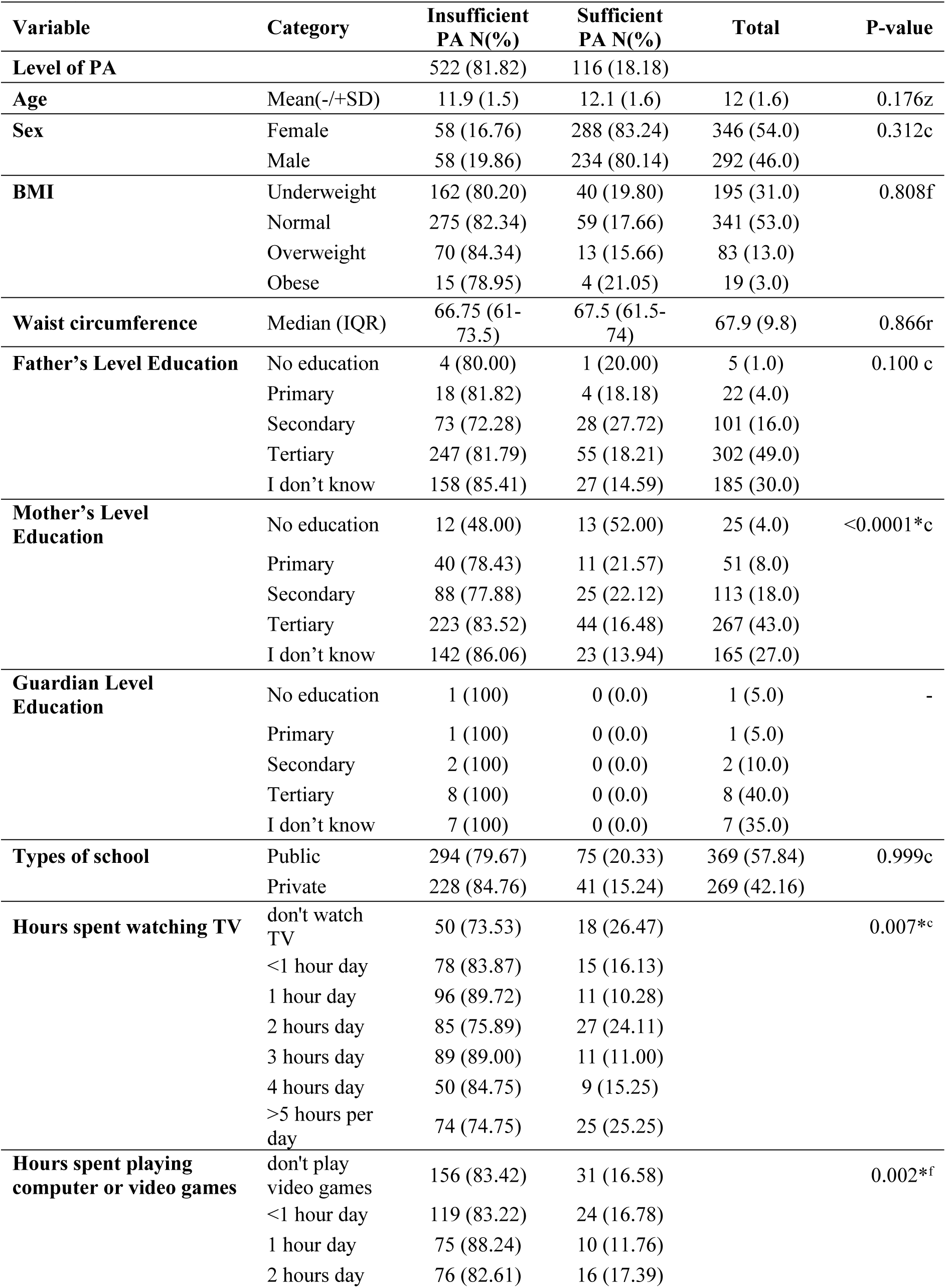
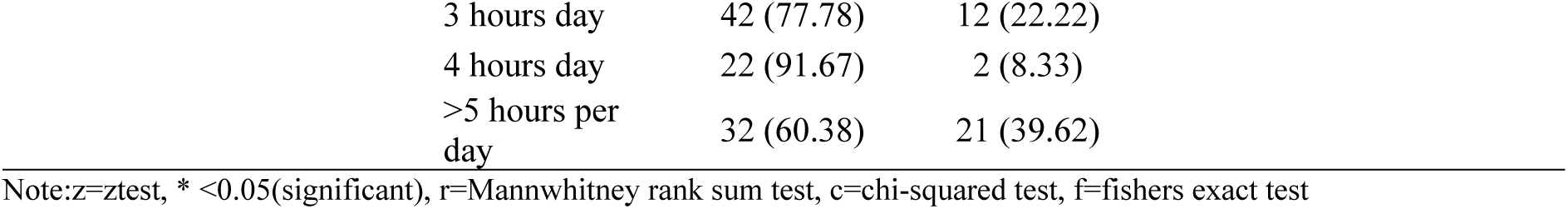
Demographic characteristics of study participants by Level of physical activity (n=638)

| Variable | Category | Insufficient<br>PA N(%) | Sufficient<br>PA N(%) | Total | P-value |
| --- | --- | --- | --- | --- | --- |
| **Level of PA** |  | 522 (81.82) | 116 (18.18) |  |  |
| **Age** | Mean(-/+SD) | 11.9 (1.5) | 12.1 (1.6) | 12 (1.6) | 0.176z |
| **Sex** | Female | 58 (16.76) | 288 (83.24) | 346 (54.0) | 0.312c |
|  | Male | 58 (19.86) | 234 (80.14) | 292 (46.0) |  |
| **BMI** | Underweight | 162 (80.20) | 40 (19.80) | 195 (31.0) | 0.808f |
|  | Normal | 275 (82.34) | 59 (17.66) | 341 (53.0) |  |
|  | Overweight | 70 (84.34) | 13 (15.66) | 83 (13.0) |  |
|  | Obese | 15 (78.95) | 4 (21.05) | 19 (3.0) |  |
| **Waist circumference** | Median (IQR) | 66.75 (61-73.5) | 67.5 (61.5-74) | 67.9 (9.8) | 0.866r |
| **Father's Level Education** | No education | 4 (80.00) | 1 (20.00) | 5 (1.0) | 0.100 c |
|  | Primary | 18 (81.82) | 4 (18.18) | 22 (4.0) |  |
|  | Secondary | 73 (72.28) | 28 (27.72) | 101 (16.0) |  |
|  | Tertiary | 247 (81.79) | 55 (18.21) | 302 (49.0) |  |
|  | I don't know | 158 (85.41) | 27 (14.59) | 185 (30.0) |  |
| **Mother's Level Education** | No education | 12 (48.00) | 13 (52.00) | 25 (4.0) | <0.0001*c |
|  | Primary | 40 (78.43) | 11 (21.57) | 51 (8.0) |  |
|  | Secondary | 88 (77.88) | 25 (22.12) | 113 (18.0) |  |
|  | Tertiary | 223 (83.52) | 44 (16.48) | 267 (43.0) |  |
|  | I don't know | 142 (86.06) | 23 (13.94) | 165 (27.0) |  |
| **Guardian Level Education** | No education | 1 (100) | 0 (0.0) | 1 (5.0) | - |
|  | Primary | 1 (100) | 0 (0.0) | 1 (5.0) |  |
|  | Secondary | 2 (100) | 0 (0.0) | 2 (10.0) |  |
|  | Tertiary | 8 (100) | 0 (0.0) | 8 (40.0) |  |
|  | I don't know | 7 (100) | 0 (0.0) | 7 (35.0) |  |
| **Types of school** | Public | 294 (79.67) | 75 (20.33) | 369 (57.84) | 0.999c |
|  | Private | 228 (84.76) | 41 (15.24) | 269 (42.16) |  |
| **Hours spent watching TV** | don't watch TV | 50 (73.53) | 18 (26.47) |  | 0.007*c |
|  | <1 hour day | 78 (83.87) | 15 (16.13) |  |  |
|  | 1 hour day | 96 (89.72) | 11 (10.28) |  |  |
|  | 2 hours day | 85 (75.89) | 27 (24.11) |  |  |
|  | 3 hours day | 89 (89.00) | 11 (11.00) |  |  |
|  | 4 hours day | 50 (84.75) | 9 (15.25) |  |  |
|  | >5 hours per day | 74 (74.75) | 25 (25.25) |  |  |
| **Hours spent playing computer or video games** | don't play video games | 156 (83.42) | 31 (16.58) |  | 0.002*f |
|  | <1 hour day | 119 (83.22) | 24 (16.78) |  |  |
|  | 1 hour day | 75 (88.24) | 10 (11.76) |  |  |
|  | 2 hours day | 76 (82.61) | 16 (17.39) |  |  |

|  |  |  |
| --- | --- | --- |
| 3 hours day | 42 (77.78) | 12 (22.22) |
| 4 hours day | 22 (91.67) | 2 (8.33) |
| >5 hours per day | 32 (60.38) | 21 (39.62) |

Note: z=ztest, \* <0.05(significant), r=Mannwhitney rank sum test, c=chi-squared test, f=fishers exact test

### Personal, Interpersonal and Environmental barriers associated with physical activity

Table 2 presents the cross-tabulations of personal, interpersonal and environmental barriers stratified by physical activity level. The only personal barriers which were associated with physical activity were feeling embarrassed and the need to shower and fix one’s hair after PA. Those who indicated that PA made them feel embarrassed had a higher rate of sufficient PA than those who were not embarrassed (18.99% vs 6.98%, p=0.03). The need to shower and fix the hair after PA was associated with a lower rate of sufficient PA compared to those who do not need to shower or do their hair after PA (12.0% vs 25.69%, p<0.001). Regarding environmental barriers, those who indicated that there is no playground or park near their home had a higher rate than those who did not (22.98% vs 15.13%, p=0.012). Children who indicated that it is not safe to walk or jog in the neighbourhood had a higher rate of sufficient PA compared to those who did not state so (21.04% vs 14.78%, p=0.041). With regards to parental support, those who were not watched by their parents’ doing PA were associated with a higher rate of sufficient PA than those who were watched (32.65% vs 16.98%, p=0.006). On peer influence, those who had peer support had a higher rate of sufficient PA compared to those without peer support (23.65% vs 16.55%, p=0.049). Individuals who did PA with their peers had a lower rate of sufficient PA than those who did not (15.79% vs 22.18%, p=0.043). Participants who were not watched by their peers doing PA were associated with a higher rate of sufficient PA than those who were watched (14.99% vs 26.90%, p=0.001). Those who were told that they are doing well in PA by their peers had a higher rate of sufficient PA than those who were not (26.76% vs 17.11%, p=0.047).

**Table 2:**
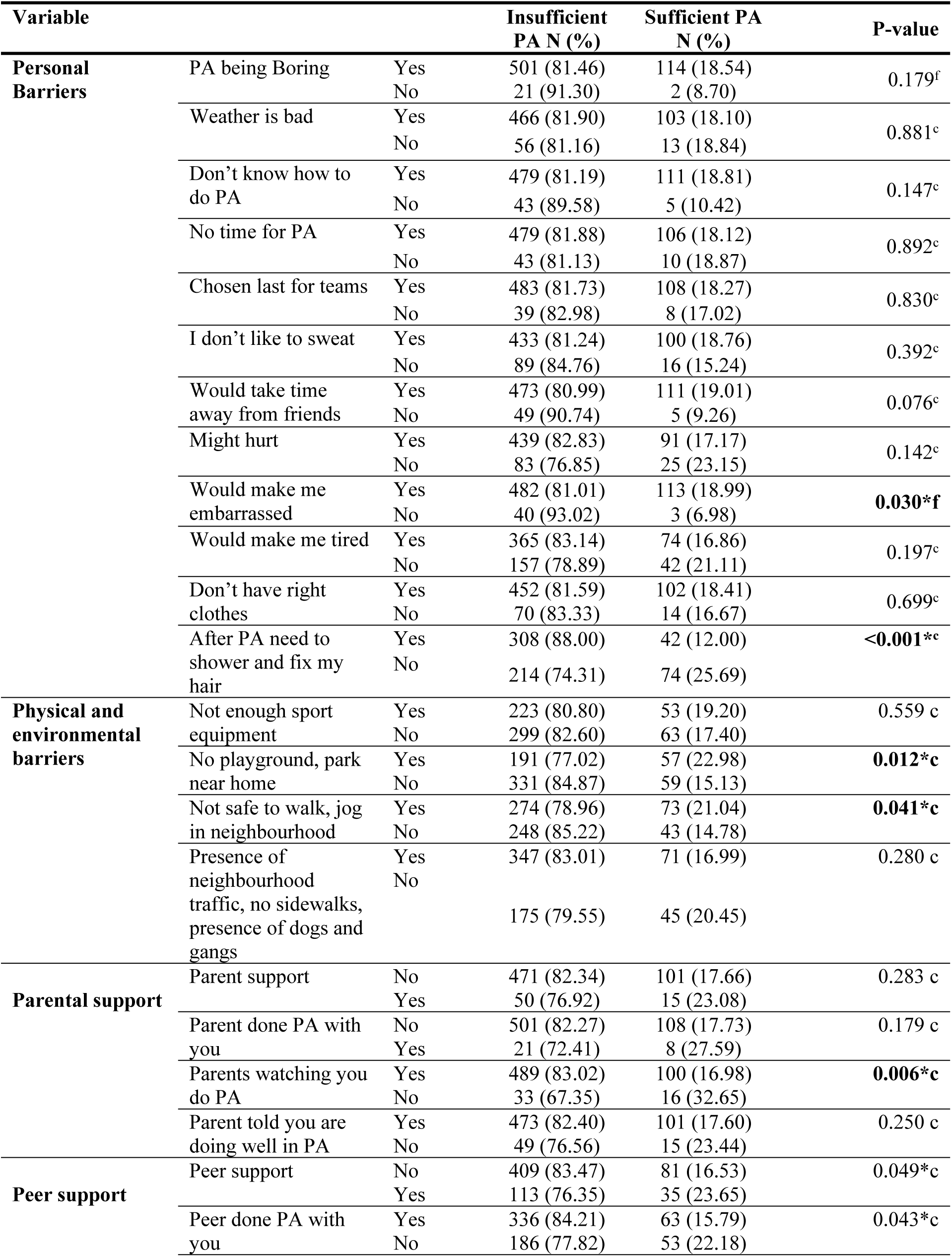
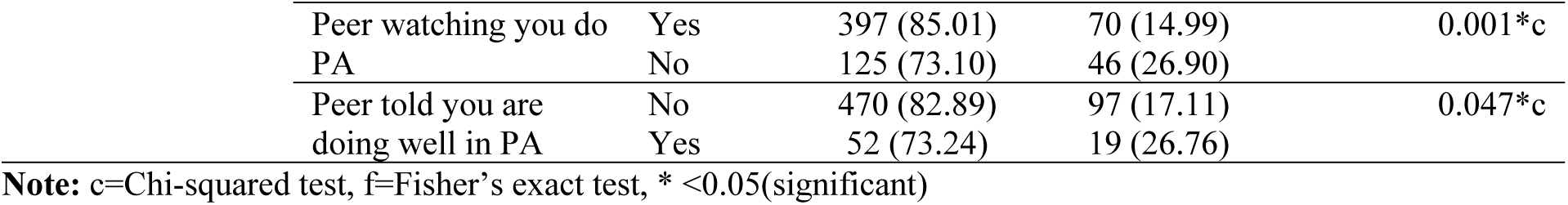
Personal, interpersonal and environmental barriers associated with physical activity.

### Risk factors associated with sufficient physical activity

Table 3 shows the multivariable binary regression analysis. Logistic regression showed that children who watched TV for one hour per day had a 69% lower likelihood of engaging in PA (aOR=0.31; 95% CI: 0.13-0.75), whereas children who watched TV for three hours daily had a 71% lower likelihood of engaging in PA (aOR=029; 95% CI: 0.12-0.73) compared to those who never watched TV daily. Children whose mothers had primary education (aOR=0.29; 95% CI: 0.09-0.88), secondary education (aOR=0.24; 95% CI: 0.09-0.64) and tertiary education (aOR=0.18; 95% CI: 0.07-0.47) were associated with a lower likelihood of having sufficient PA compared to those whose mothers had no education. Children who were never bothered by the need to shower and fix their hair had a 94% higher likelihood of having sufficient PA compared to those who were bothered (aOR=1.94; 95% CI: 1.21–3.11).

**Table 3:** Multivariable binary logistic regression for factors associated with sufficient physical activity.

| Variable | Category | AOR | AOR Percentage | p-value | 95% CI |  |
| --- | --- | --- | --- | --- | --- | --- |
| **Mother LE** | No education | ref |  | ref | ref |  |
|  | primary education | 0.29 | 71 | **0.03\*** | 0.09 | 0.88 |
|  | secondary education | 0.24 | 76 | **0.005\*** | 0.09 | 0.64 |
|  | tertiary education | 0.18 | 82 | **<0.001\*** | 0.07 | 0.47 |
|  | I don't know | 0.18 | 82 | **0.001\*** | 0.07 | 0.48 |
| **Hours watching TV** | Not watch TV | ref |  |  |  |  |
|  | <1 hour day | 0.62 | 38 | 0.265 | 0.27 | 1.44 |
|  | 1 hour day | 0.31 | 69 | **0.01\*** | 0.13 | 0.75 |
|  | 2 hours day | 0.91 | 9 | 0.805 | 0.42 | 1.96 |
|  | 3 hours day | 0.29 | 71 | **0.008\*** | 0.12 | 0.73 |
|  | 4 hours day | 0.53 | 47 | 0.196 | 0.20 | 1.39 |
|  | >5 hours per day | 0.76 | 24 | 0.505 | 0.34 | 1.69 |
| **Peer support- played with you** | No | ref |  |  |  |  |
|  | Yes | 1.15 | 15 | 0.603 | 0.67 | 1.97 |
| **Peer support-watched you play** | No | ref |  |  |  |  |
|  | Yes | 1.62 | 62 | 0.097 | 0.92 | 2.85 |
| **Peer support-told doing well** | No | ref |  |  |  |  |
|  | Yes | 1.42 | 42 | 0.293 | 0.74 | 2.73 |
| **Personal barrier-shower, fix hair** | Yes | ref |  |  |  |  |
|  | No | 1.94 | 94 | **0.006\*** | 1.21 | 3.11 |
| **Personal barrier-no time for PA** | Yes | ref |  |  |  |  |
|  | No | 0.95 | 95 | 0.829 | 0.58 | 1.54 |
| **Environmental barrier-bad weather** | Yes | ref |  |  |  |  |
|  | No | 0.83 | 17 | 0.447 | 0.52 | 1.34 |

Note: AOR=adjusted odds ratio; \* significant $p<0.05$ ; ref=reference group

## Discussion

This study aimed to assess the physical activity levels and associated factors among upper primary school children in Lusaka. It was revealed that over three-quarters of the respondents were insufficiently active and this was significantly associated with excessive screen time, high maternal education, lack of recreational spaces, and limited social support.

The study showed that less than one-fifth of the participants were sufficiently active. The levels of sufficient PA in our study were higher than some sub-Saharan counties namely, Sudan (11%) and Mauritania (14%) and some Asian countries like Cambodia (9%) and the Philippines (8.7%) [3]. However, our findings were lower than countries with strong PA policies, such as Finland (59.6%), Canada (58.6%) and the U.S. (57.3%) [3]. This raises the risk of obesity, cardiometabolic illness, poor bone health, and mental health issues, which is a serious public health concern among upper primary school children in Lusaka, Zambia.

Our study revealed that students in public schools were more physically active than their counterparts in private schools, although not significant. This disparity may stem from the recent proliferation of private schools in Zambia, which are often situated in residential areas with limited space for recreational facilities like playgrounds. This finding is consistent with a Brazilian study that also reported higher physical inactivity in private versus public schools [21]. Furthermore, the de-prioritisation of Physical Education (PE) in favour of subjects like Mathematics and Science represents a significant barrier, a conclusion supported by research in both Zambia and Ghana [22,23]. Compounding these issues, excessive screen time was identified as another key factor in reducing learners’ participation in physical activities.

In the current study, children of mothers with higher educational attainment were less active than those of less-educated mothers. This may reflect lower sedentary time and greater reliance on active transport among children of less-educated mothers, who often have reduced access to digital devices and private vehicles. Evidence on the effect of maternal education on childhood physical activity is mixed, with socioeconomic context often shaping activity type [24,25].

Parental and peer support were also significant motivators. Children with supportive parents were more likely to engage in PA, while excessive parental control discouraged it. This supports literature showing that parental support positively influences children’s PA and reduces sedentary time [26,27]. Peer encouragement also increased PA participation, reinforcing the importance of social interactions in fostering active lifestyles.

A strong association was observed between PA levels and body weight. Overweight children were predominantly inactive (84%), and obesity negatively affected physical performance and confidence, discouraging participation. These findings align with Casonatto et al. [28], who reported that higher BMI and abdominal obesity adversely impacted physical performance, including flexibility and trunk reachability.

Time constraints were another major barrier, with 82% of inactive children citing lack of time as a reason for insufficient PA. Similar patterns have been reported globally, highlighting the importance of school-based interventions that incorporate PA into daily routines [29]. Psychological barriers, such as embarrassment and post-activity grooming concerns, particularly among girls, also discouraged participation. Hair maintenance concerns were significant in this study and have similarly been cited as barriers among African-American women, limiting exercise [30].

Environmental factors such as bad weather and lack of recreational spaces also pose risks to the reduction of PA levels. Chaabane et al. [31] similarly reported that extreme weather conditions reduced PA in several Middle East and North African countries. In this study, limited access to playgrounds and safe spaces further discouraged PA, while poor neighbourhood conditions, including traffic congestion and crime, were additional barriers. This finding aligns with literature emphasising that the absence of convenient recreational facilities such as parks, open spaces, gyms, and sports clubs contributes to inactivity [32].

Policy implications from these findings are clear. Schools should integrate structured PA programs, and campaigns should raise awareness on reducing screen time, addressing psychological barriers, and fostering motivation. Urban planning must prioritise safe and accessible parks, sidewalks, and pedestrian-friendly infrastructure to encourage PA. Strengthening parental and peer support systems is also essential in promoting PA among school-aged children.

### Strengths and Limitations

The current study has a number of strengths. Firstly, a large dataset representing the school-going children from all 10 zones of Lusaka Urban District was used. Secondly, a multivariable binary logistic regression model was used to determine the relationship between PA and the predictor variables of PA, which controlled for confounders. Thirdly, the study established the levels of PA and the factors that influence PA among children, which may be a basis to call for concerted effort and immediate action to improve PA among relevant key stakeholders.

The study had some limiting factors that are worth mentioning. Firstly, the study was carried out in Lusaka Urban District and therefore, may not be generalised to the entire school children population in Zambia. Secondly, the physical activity self-reported surveys may be subject to recall bias and therefore must be construed within that context. Thirdly, the study design is cross-sectional; this type of study does not allow causal relationships to be established. Finally, the sampling process for private schools was purposeful rather than random; this process is inherently susceptible to researcher bias.

## Conclusion

This study shows that 82% of children did not meet PA guidelines, putting them at risk of non-communicable diseases (NCDs). Barriers included excessive screen time, low maternal education, lack of recreational spaces, and limited social support. A multi-sectoral approach is required, including mandatory PA sessions and PE programs in schools, parental involvement to promote active play and reduce screen time, and urban planning to provide safer environments. Promoting PA among Zambian schoolchildren is vital for long-term public health. Evidence-based interventions will foster healthier lifestyles and reduce the future burden of NCDs.

## Data Availability

Data sets may be available upon request from the corresponding author.

## Acknowledgements

The authors wish to thank the Department of Physiotherapy at the University of Zambia in the School of Health Sciences for their contribution and guidance, and also the Ministry of Education, the head teachers from various schools, the parents and the school children for their support and valuable time throughout the study.

## Ethics approval and consent to participate

Ethical approval to conduct the study was granted by the University of Zambia Health Sciences Research Ethics Committee (UNZAHSREC) under reference number: IRB no: 00011000, IORG no: 0009227, FWA no: 00026270 and protocol ID: 20203101012. Further permission to conduct the study was sought from the head teachers in the various schools, while assent was obtained from the parents and guardians of the children.

